# A Model to Introduce Medical Students to the Use of Artificial Intelligence and Genomics for Precision Medicine

**DOI:** 10.1101/2021.05.13.21255493

**Authors:** Philip O. Alderson, Maureen J. Donlin, Lynda A. Morrison

**Affiliations:** Saint Louis University School of Medicine

## Abstract

**Objective:** Despite the significant medical impact of artificial intelligence (AI) in healthcare, emergence of AI-related topics in medical curricula has been slow. The authors sought to introduce pre-clinical students to the importance of AI methodologies and medical applications using modular short courses focused on active learning with precision medicine as a primary use case.

**Materials and Methods:** A short elective course was designed to introduce first-year students to how various bioinformatic and AI-related processes work and how they help classify medical data, facilitate genomic analysis and predict clinical outcomes. The course covers gene sequencing and variants, neural networks, natural language processing, medical computer vision and the limitations and ethical concerns related to use of AI in precision medicine. Online content serves as major source material. After a faculty-led introduction, sessions focus on teams of students who present course content to one another and lead discussions with faculty guidance. A related short AI course focused on gene variants was given to the entire second-year class.

**Results:** The elective course has been taken by 74 first- year students over 8 consecutive semesters (2017-2021). The course achieved average satisfaction scores of 4.4/5.0 (n = 13) when the active learning approach became dominant in 2018. Students were able to describe accurately how bioinformatics and AI make personalized medicine possible. Students also did well on the gene variants exercise given to the entire second year class (2018), but the full class short AI course was not continued in subsequent years. Students have created a school-approved interest group in medical AI.

**Conclusions:** This experience shows that AI-related materials can be sustainably introduced into pre- clinical medical education with precision medicine as the primary use case. This modular course design and content could be adapted easily for educational use in medical subspecialties and other health professions.

## Objective

In recent years, AI methodologies have allowed a host of medical applications to achieve sufficient accuracy, speed and affordability to become clinically useful and practical ^1,2^. In somewhat similar ways, clinical genomics as represented in personalized/precision medicine also has become practical. Whole genomes now can be sequenced within 48 hours at a cost approximating $1,000 ^3^. This progress has been made possible by machine automation, advances in computational bioinformatics and application of deep learning (DL) AI technologies to genome sequencing. Pre-designed hybridization gene arrays also are available, are much less expensive and are becoming important in direct to consumer marketing associated with precision medicine ^4^. AI in the form of deep neural networks also is being applied with increasing frequency to a variety of healthcare issues including medical image classification, natural language processing (NLP) and interrogation of medical databases including electronic health records (EHRs), mobile health diagnostics and telehealth. Such applications coupled with improved gene sequencing and gene arrays are making personalized/precision medicine (PM) a reality ^5^.

Despite the rapid rate at which big data approaches are developing and predictions that PM will transform medical care ^2^ the inclusion of AI-related machine learning (ML) and related genomics and PM in medical curricula has been slow ^6,7^. Readily understandable, concept-based medical teaching materials are not widely available. Because AI and PM developments have occurred rapidly in the past 8 to 10 years, many medical faculty are not comfortable teaching AI / PM and students are not learning how PM can benefit patients. The authors sought, therefore, to demonstrate that AI-facilitated PM could be presented in modular short course formats in an accessible way to get students engaged with the rapidly evolving impact of AI in medicine.

## Materials and Methods

Introductory courses were developed that integrate AI principles with PM applications. First, a standard 6-session elective for first-year students was introduced in the Fall term of the 2017- 18 academic year. The elective was titled, “Understanding Artificial Intelligence for Precision Medicine.”

By the end of the course students should be able to:

1. Describe the potentials of artificial intelligence (AI) in precision medicine (PM).
2. Understand clinically relevant aspects of AI in PM.
3. Explain how big data science is being used to make PM possible.
4. Use appropriate websites to explore AI- related PM topics.
5. Articulate limitations of medical AI in PM.
6. Discuss the ethics of using these tools to make medical decisions.

An active learning format was used in this course. At the end of an opening session in which faculty previewed course content, small teams of students were created. Students were told that the teams would remain together throughout the course. Faculty assigned one or more relevant topics at the end of each session and asked student teams to report back to the other teams at the next session. Teams were asked to organize their report backs in whatever way they think would best inform the other teams. They were told that they may select their own presentation format, e.g., PowerPoint, blackboard, demonstration, and that all students on each team should participate in the presentation if possible. The classrooms that were used always had computer- linked projection. All students were asked to bring their own laptops to each session to facilitate in-class website exploration. Student mastery of material was reflected by the quality of report back assignments and in-class participation. An impression of these activities was recorded by the lead faculty member during the sessions and used to aid in submission of a Saint Louis University (SLU)-standard qualitative (pass/fail) student performance summary at the end of the course.

The exact sequence of topics in the sessions varied a bit from semester to semester but each session contained AI topics and PM applications. Faculty guidance was given as needed to show how the AI and PM topics were related. Topics with straight forward AI-PM links were assigned early in the semester. Later sessions focused on more complicated material. The following paragraphs provide the methods and rationales for this structure. Additional details are provided in the table immediately below and in the Course Syllabus (Appendix).

### Highlights of An Integrated Short Course in AI for Precision Medicine

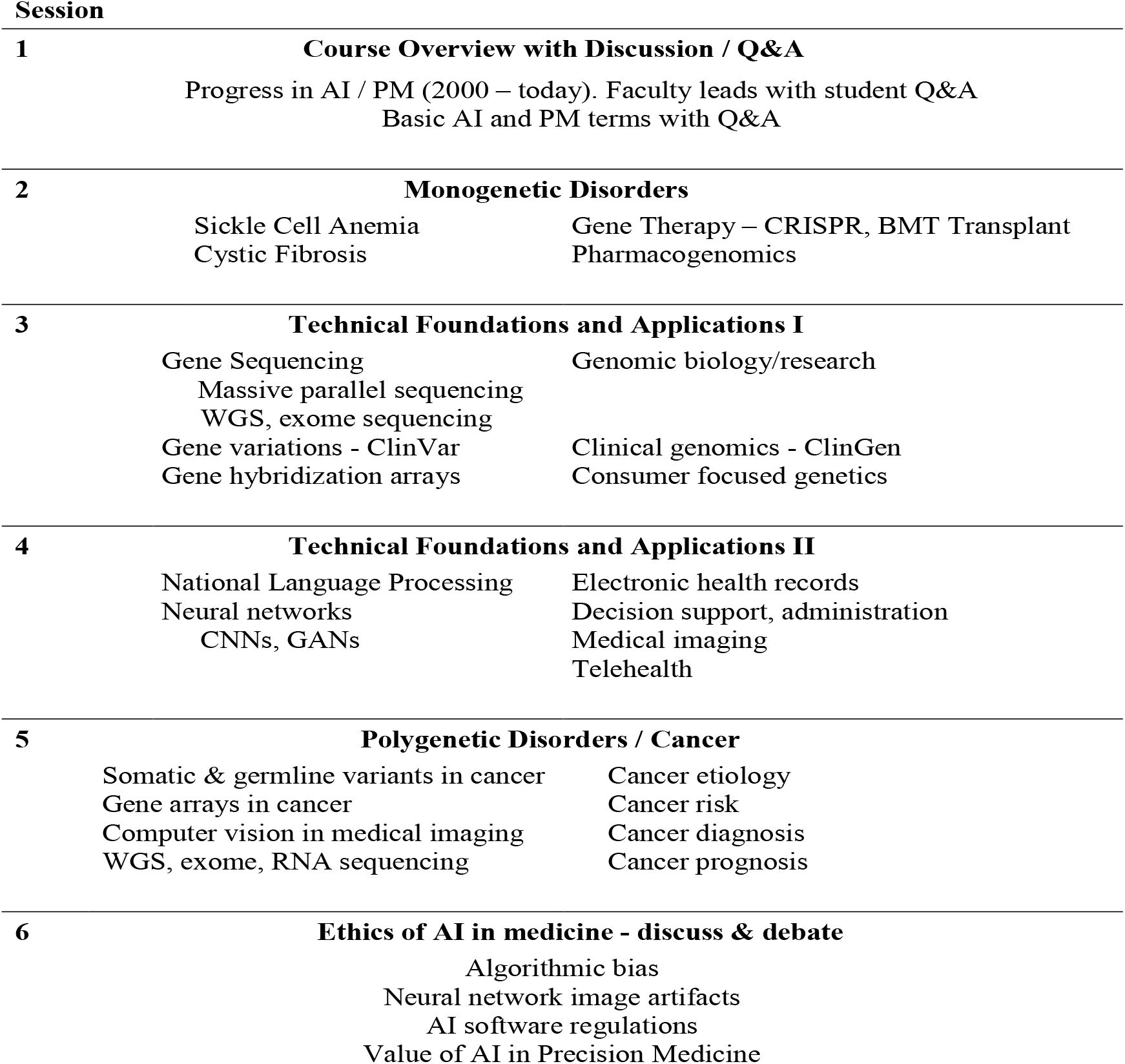

The first session always involved some discussion of logistics and getting to know one another. It also involved a PowerPoint-guided overview of course content presented by the faculty leader with periodic questions to students to create active involvement. Definitions of basic AI/bioinformatic terms were introduced and discussed with examples, e.g., that an algorithm is the set of line-by-line software instructions in computer code that guide the program’s actions. Additional terms such as narrow AI, machine learning (ML), supervised and unsupervised ML also were introduced. Examples of the importance of mathematics and statistics in AI and the importance of computer coding in bringing AI algorithms to life were provided by faculty. The faculty leader and students briefly discussed the mathematical basis of machine learning and how receiver operating characteristic curves (ROC) are used to compare AI performances. The course focus, however, was on conceptual content about how AI/bioinformatics enabled PM rather than the mathematical/statistical basis of AI.

The second session, which featured the first report-backs, focused on well-studied single gene to phenotype relationships found in Mendelian disorders like sickle cell anemia (SCA), cystic fibrosis (CF) and familial colonic polyposis. These disorders also provided the opportunity to discuss recent progress such as CRISPR-based gene therapy for SCA and pharmacogenomic progress in treating CF. All student groups were directed to the Hopkins-curated On-line Mendelian Inheritance in Man (www.OMIM.org) site ^8^ and to the gene cards site (www.genecards.org) to learn about these disorders and report back to their classmates.

In another session students were asked to report back on the basics of gene sequencing and how gene variants are defined. A student team was asked to describe bioinformatic approaches to massive parallel gene sequencing ^9^, including use of the genome reference standard and limitations in massive parallel approaches for whole genome sequencing (WGS) and exome sequencing. Students also defined gene hybridization arrays and compared them to WGS. To get at the clinical problem of gene variants, especially variants of uncertain significance, a student team reviewed the NIH/NLM site ClinVar ^10^. Students were asked to demonstrate to their classmates how gene variants are reported, reviewed and ranked in ClinVar. Another student group reported on the NIH Clinical Genome (ClinGen) site^11^ to show how gene variants are determined to cause a clinical disorder and to be actionable if FDA-approved medications are available to treat the disorder.

In another foundational session, student teams were asked to report to their classmates on the basics of medical databases and natural language processing (NLP) ^12^. Use of NLP in EHRs and basics of EHRs as transactional and analytic databases also were reported in this session. In semesters when EHR experts were available to contribute to the sessions, they led the discussions of EHR functionality and taught students to access a simulated EHR program (EPIC hyperspace playground) to explore simulated transaction cases. Students did some explorations with expert tutelage during the class session and were encouraged to pursue additional simulated EHR interactions outside the classroom.

In another session, a student team provided a report on neural networks with attention to recent advances that have led to widespread research and clinical utilization of deep neural networks ^13^. A report back on deep neural networks configured for computer vision, i.e., convolutional neural networks (CNNs), was provided in this session. This session included faculty-provided examples of computer vision in face recognition and in self-driving vehicles. Students reported on the use of CNNs in various types of medical imaging to set the stage for a report-back on computer vision applications in cancer diagnosis in a later session.

A late-stage session, typically the second to last session of the course, focused on cancer as the exemplar of complex polygenic disorders. Polygenic risk scoring was briefly addressed, typically by the faculty member. A student team reported on The Cancer Genome Atlas ^14^ (TCGA) during this session. In the same session, another student team typically would report back on published clinical cases or studies in which the combination of genomics with data science was instrumental in clinical management of cancer ^15^. This session also included a report back on how gene arrays are being used in precision oncology ^16,17^ and how their clinical utility compares to exome or whole genome analysis. This typically busy session also included a student report on how CNNs are being used for classification of radiologic, pathologic and dermatologic image findings in patients being evaluated for cancer.

In the final session of the course, students presented and discussed limitations and potential biases of AI technologies and algorithms, including artifacts and mis-diagnosis potentials in CNN-based medical imaging. This set the stage for open discussion of the ethics of using AI approaches to make clinical decisions. Open discussion and debate of AI/bioinformatic-enabled PM typically concluded the final session.

To extend the materials provided in the elective course and to focus on the importance of gene variants, a 6-member ad hoc faculty committee that included the three co-authors of this paper arranged for a 3-session MLDS course to be presented to the entire second year class as part of the regular second year curriculum in 2018-19. A lecture on statistical methods used in ML along with overviews of AI methods and clinical applications started the course, but the focus of the course was on understanding gene variants. This included a student exercise involving characterization and validation of “unknowns” created from the ClinVar database. This pass/fail exercise was designed for students to be able to:

1. Evaluate the medical significance of genome variants using the ClinVar database.
2. Examine the gene in which a variant is found and the potential impact of the variation on the encoded protein.
3. Explain the potential medical impact of specific variants to a patient.

A set of 152 single nucleotide polymorphisms (SNPs) variants covering 22 different genes that had a review status of at least 2 stars was selected. This rating indicated that these variants had multiple submitters for the same variant and no conflict. The students were divided into 30 groups of 6 students each. Six variants were assigned to each group, with at least one of those having a pathogenic outcome. Background material was provided on the prevalence of human variation, the contribution of genome sequencing projects to identification of variants and the development of ClinVar as a publicly accessible database of variants and their clinical significance. After an in-class tutorial, the students were to identify the genes in which their assigned variants are found and the disease, pathology or risk factor associated with each variant, if applicable. Finally, students were asked to provide a description of how they would communicate the potential clinical outcome of a gene variant to a patient. This was done for each assigned variant, whether it had a disease pathology or was considered benign. The students were to understand the following key points:

1. A variant is not the same as a pathological mutation.
2. Every genome possesses ∼1 million variations relative to the “reference” human genome.
3. Within the same gene, some variants can contribute to pathology or risk factor while others are benign.
4. Very few SNP variants are clinically actionable.

This exercise was completed by the entire second year class of 180 students. To ensure that the amount of effort required to work up each variant was equitable, only SNPs that caused a missense mutation were used. This reduced the number of variants that had severe pathologies but reinforced the concept that a variant does not equal a deleterious mutation. The student group reports were filled out online and the 30 student group reports were divided equally among the six faculty on the course ad hoc committee for pass/fail grading. This pass/fail assessment was independent of the assessment of students in the elective course.

## Results

The elective course has been taken by a total of 74 first-year students over the 8 semesters in which the course has been given. Student teams in both courses proved adept at active learning exercises. In the elective, student teams provided well organized and thorough report-backs on AI techniques and AI-facilitated precision medicine topics that increased in complexity as the course progressed. Depending on the elective class size, the teams had as many as five students. One semester when only 6 students took the elective, individual students completed the feedback assignments quite adequately. In the whole class gene variants exercise, all teams completed the gene variant unknowns accurately and on time. These active learning exercises were popular and effective with the students and the shared grading approach showed how an active learning exercise could be easily documented for a class of 180 students. The popularity of active learning was reflected in the student ratings of both the elective and whole class short courses.

In the elective, faculty lectures with PowerPoint slides were a major part of the teaching approach in the first year. SLU-standard feedback combined for both semesters was provided at year-end and indicated that students preferred active learning to lectures. Accordingly, student presentations and discussions of assigned website materials became the dominant learning approach in the following year. This approach was responsible for an increase in student satisfaction ratings. In the first year, the overall student satisfaction rating for the elective was 3.9 (n = 18 voting) on a 5-point scale. In the second year, overall student satisfaction with the quality of the elective grew to 4.4 (n = 13) and the rating for the quality of feedback to students increased from 4.0 to 4.5. As a result, the active learning approach remained the dominant educational approach in subsequent years.

The student responses in the gene variants course were similar. A group of eight students, who had signed up at the beginning of the semester to provide feedback on the course as per SLU policies, reviewed the ClinVar exercise with the faculty leader. Overall, they liked the gene variants exercises and thought learning about ClinVar would be useful in their careers. They said they would prefer better integration of the gene variants exercise with clinically related course materials, such as the year-2 cardiovascular module. They felt that the AI lecture that opened the gene variants course would be better if paired with more detailed clinical material and stated that it was better suited to the elective course format. Because of stresses on time slots in the regular year-2 curriculum, the AI /gene variants course was not repeated in subsequent years. As a result, attention to ClinVar was increased in the elective course but it was not as extensive as the full class gene variant exercise.

Because of the positive reception to and perceived importance of medical student education in AI, there have been related developments. Invitations from the dean’s office have come each year to summarize medically relevant AI during the senior year capstone course, providing a way to deliver AI information and recent developments to students who have not taken the elective. These presentations have been well received. Second-year students who took the PM elective in 2017-2018 created a formal Interest Group in Precision Medicine and invited local and visiting lecturers. The interest group continues but presentations were cancelled during the COVID crisis and have not yet resumed. During the spring 2020 elective when COVID arose, SLU-wide polices resulted in the final elective session being cancelled. To help compensate, an email was sent to the students with reading materials addressing ethics of AI in healthcare. During the fall semester of 2020 and spring semester of 2021, all classes were held and were in person following COVID protocols.

## Discussion

Our experiences demonstrate that a faculty-led short course emphasizing scientific concepts underlying the data science and genomics in personalized medicine (PM) can succeed in conveying these concepts and achieving learning satisfaction and engagement in medical students. To our knowledge, this paper reports the only successful AI introductory course model that has been sustained in a medical school over a period of years. We believe that an understanding of PM concepts is important in preparing physicians and other health professionals to make appropriate clinical and ethical decisions about future applications of AI in healthcare. Our experiences also suggest that concurrently teaching AI principles with AI-related aspects of clinical genomics and other practical medical applications is an excellent way to demonstrate the importance and interdependence of these scientific disciplines on precision medicine.

Finding effective teaching materials and staying current turned out to be among the most time consuming and rewarding aspects of developing and sustaining the elective course. Website materials are an important source of current AI developments, but these materials must be carefully curated. Many sites are not peer-reviewed and some are frankly commercial. Web-based materials that originate from data science / engineering groups, though excellent in many ways, often progress quickly to equation-based mathematical and statistical explanations. We sought and used more concept-based materials, focusing on those that had medical applications. To stay current, new materials had to be added nearly every semester.

Interdisciplinary collaborative teaching helped expand the scope of the AI-PM message. The course described in this manuscript could not have been delivered without the collaboration of physicians, bioinformatic scientists and EHR experts. Broader collaborations involving subspecialty physicians, biomedical engineers, non-MD health professionals and educators could help create and sustain high-quality medical AI education to an even wider audience. The time is right for this to happen.

AI technologies are making a major impact on healthcare. In 2020, a Harvard webinar ^18^ described digital health technologies as a “pillar” upon which healthcare would be built and showed how growth was accelerating in the midst of the COVID pandemic. On another common healthcare front, a recent paper from Lincoln et al ^19^ reported that tumor tissue genome sequencing coupled with blood tests for germline mutations reveals higher than expected impacts of inherited contributions to a variety of common cancers. Such developments coupled with the growing number of gene arrays now being marketed directly to consumers ^4^ exemplify the importance of familiarizing medical students with the basics behind AI and genomics in precision medicine ^16,17^.

According to the National Academy of Medicine ^1^, physicians should know how AI works, when to use it and its limitations. Wartman and Combs ^20^ indicate that the educational move toward AI in medicine means more than simply updating medical curricula. It is a transition from traditional memory-based learning to educational approaches that teach students how to find and analyze the information they need. In 2020, when posted medical knowledge is said to double every 73 days ^21^, healthcare professionals need to understand how to use AI to find, critique and assimilate the data they need. Despite these needs, AI and related personalized medicine have not been well covered in medical education ^6,7^. Shortliffe opined more than 10 years ago ^22^ that this was because extant medical curricula were overly full and because there was resistance to adding new topics. Though the many positive developments in the past 10 years make clear the importance of AI and PM, curricular constraints do not seem to have changed.

Park et al ^23^ wrote in 2019 that the time is right for electives to be used to address the AI education gap in medical schools. The AI elective reported herein began in 2017 and continues successfully eight consecutive semesters later. This strongly supports that short modular AI-PM courses with active learning formats can succeed and be sustained. It is our hope that concept-based modular AI-PM courses of the type reported in the current paper will be further developed, will become part of standard medical curricula and be a format for extending AI-PM education to GME programs, nursing, allied health and other health professions.

## Supporting information

Equator

## Data Availability

Data are available in the corresponding author's files and in the Oasis medical administration website for student performance.

## APPENDIX Syllabus: MEDC-109 Spring Term 2021

1. Course: Understanding Artificial Intelligence for Precision Medicine
2. Leader: Philip O. Alderson, M.D. Student Affairs, Caroline 100
3. Learning Objectives: By the end of the course students should be able to:
  - Describe the potentials of Artificial Intelligence (AI) and Precision Medicine (PM)
  - Understand clinically relevant aspects of AI and PM
  - Explain how big data science is being used to make PM possible
  - Use appropriate websites to explore AI-related PM topics
  - Articulate limitations of medical AI and PM.
  - Discuss the ethics of using these tools to make medical decisions
4-6. Requirements / Grading / Attendance: This is a Pass/Fail course that requires attendance, preparation for and active participation in classroom sessions. These sessions will begin at 1:00 p.m. in xxxxx on the dates specified by the School of Medicine Curriculum Committee. For the spring term in 2021 the dates are xxxxx. Please bring a laptop or other computer to each session because we will be using various websites as course materials during the sessions.
7. The SLU policy on academic honesty and integrity applies to this elective course and can be found xxxx.
8. Regarding disability accommodations, the SLU policy applies to this course and can be found at xxxxx. Title IX can be found at xxxxx.
9. The spring 2021 sessions and all activities associated with the course will be governed by SLU’s policies related to COVID-19 precautions. Masks are required at all group sessions including small group meetings of students to prepare for a coming full group session. Social distancing and masks should be used in all sessions. If you feel ill or have had contact with someone who has tested positive for COVID, please notify the University and go into protective quarantine until you can be tested and treated, as required.
10. Background and Course Summary:

The first draft of the human genome was published in 2001 and the full sequenced human genome was declared complete and published in 2003. It took years to complete at costs estimated to range between 150 million and over 1 billion dollars. Today the human genome can be sequenced within several days for approximately $1,000. Other gene tests such as exome sequencing and genotyping arrays cost only hundreds of dollars. The accessibility of these tools provides an enormous array of new research and clinical opportunities that are beginning to be applied in clinical medicine.

The current efficiency of gene sequencing depends on desktop machines that “read” many gene segments simultaneously. This is called massive parallel sequencing or next generation sequencing. The machines reconstruct gene sequences using built-in powerful computers. These computations are guided by line-by- line instructions in computer code called algorithms.

The multidisciplinary field related to the analysis and interpretation of genomic sequencing and many other forms of biological data is called bioinformatics and has existed for decades. Progress in the area known as machine learning (ML) accelerated greatly in 2012 when an approach known as “deep” machine learning (DL) began to achieve wide recognition. Subsequently, research had shown that DL can be applied successfully to a myriad of medical and non-medical challenges. In this course, we will explore DL applications in medicine. We will learn how computation approaches called computer vision are used to analyze and classify medical images in radiology, anatomic pathology, dermatology and ophthalmology, among others. We will learn how computers use DL approaches to interpret written digital text and spoken words in what is known as natural language processing (NLP) and how NLP can be linked to computer vision to produce medical reports and to interrogate voluminous digital databases of many types including electronic health records (EHRs).

The application of these powerful bioinformatic deep learning tools to genomic data, personal clinical data and a myriad of normative clinical databases is allowing the dream of precision medicine/precision health to move toward reality. In the course we will explore new clinical research in precision medicine and link these developments to classical Mendelian genetics as well as to disease classification and outcome predictions in polygenic disorders like cancer. We also will explore weaknesses inherent in these big data approaches and discuss/debate the many ethical issues that arise as these computational approaches are applied in clinical medicine.

Because the field of precision medicine is evolving rapidly, the reference materials for this course are upgraded frequently and include websites and materials from science blogs. Some of you may discover other useful sites during your explorations of materials for class presentation. Please share such new sites with us. Prior to each session I will contact the class via email to review the subject matter and assignments for the next full group session. A session-by-session outline of the course follows.

### Highlights of An Integrated Short Course in AI for Precision Medicine

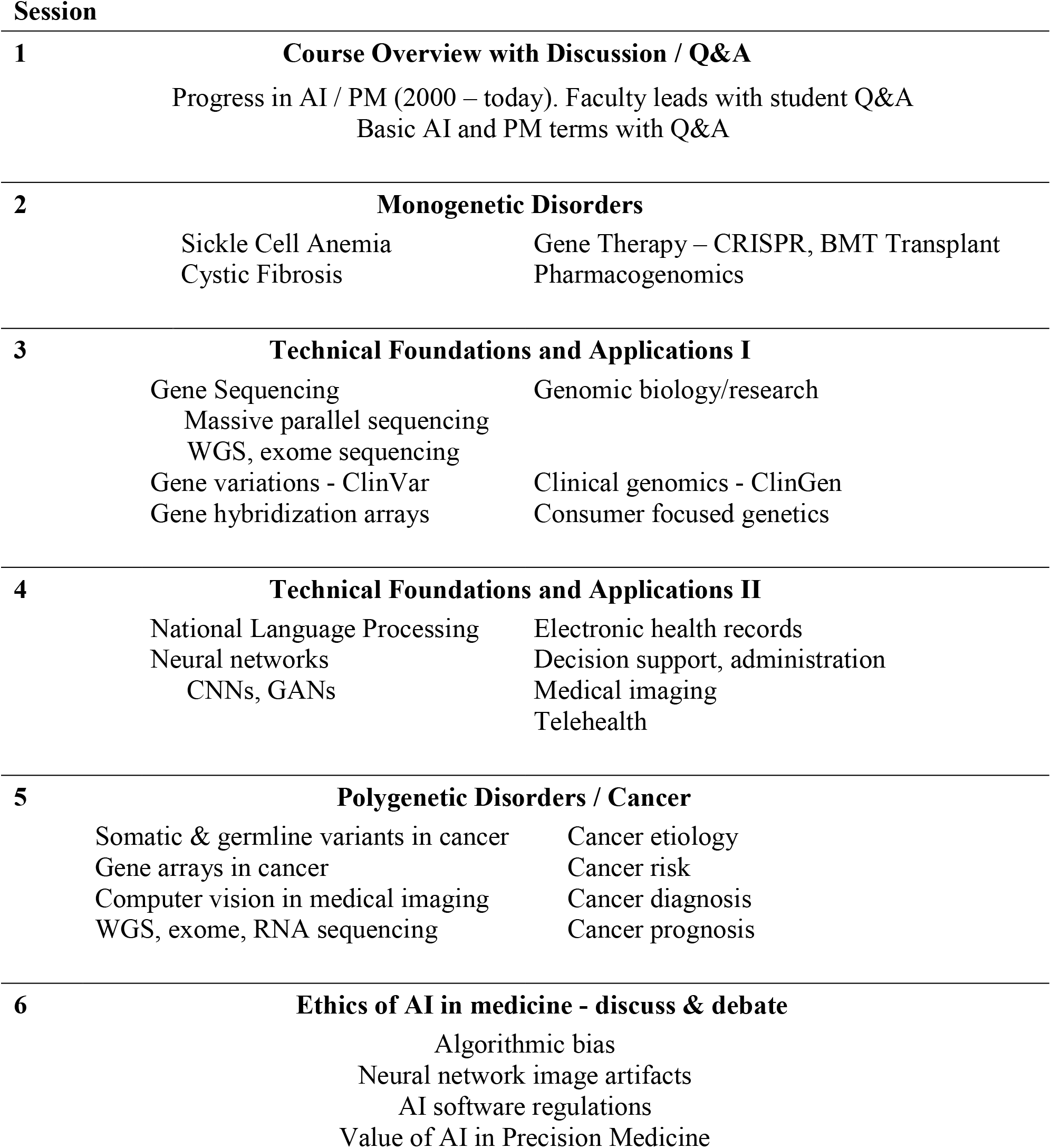

## References

1. Matheny ME, Whicher D, Thadaney S: Artificial intelligence in health care – A report from the National Academy of Medicine. JAMA 2020; 323(2): 509–510.

2. Hinton G: Deep learning - a technology with the potential to transform health care. JAMA 2018; 320(11): 102–1102.

3. Hayden EC: The $1,000 genome. Nature 2014; 507(3): 294–295.

4. Allyse MA, Robinson DH, Ferber MJ, Sharp RR: Direct-to-consumer testing 2.0: Emerging models of direct-to-consumer genetic testing. Mayo Clin Proc 2018; 93(1): 113–120.

5. Green, E: From the human genome project to precision medicine: A journey to advance human health. https://www.youtube.com/watch?v=C4dT3JWXCDI. Published July 17, 2018. Accessed various times. Most recent access March 27, 2021.

6. Kolachalama VB, Priya SG: Machine learning and medical education. NPJ Digit Med 2018; 1: 53–55.

7. Plunkett-Rondeau J, Hyland K, Dasgupta S: Training physicians in the era of genomic medicine: Trends in undergraduate medical education. Genet Med 2015; 17: 927–934.

8. Online Mendelian Inheritance in Man, McKusick-Nathans Institute of Genetic Medicine, Johns Hopkins University School of Medicine. https://www.omim.org/. Published 1987 and updated frequently. Accessed various times. Most recent access November 10, 2020.

9. Zhang J, Chiodini R, Badr A, Zhang G: The impact of next-generation sequencing on genomics, J Genet Genomics 2011; 38 (3): 95–109.

10. Clinical Variants (ClinVar), National Center for Biotechnology Information, U.S. National Library of Medicine, https://www.ncbi.nlm.nih.gov/clinvar. Published April 2013 and updated frequently. Accessed various times. Most recent access September 23, 2020.

11. Clinical Genomics Resource (ClinGen) National Center for Biotechnology Information, U.S. National Library of Medicine. https://www.clinicalgenome.org. Published 2013 and updated frequently. Accessed various times. Most recent access March 25, 2021.

12. Naik A: A contemporary introduction to natural language processing in 2020. https://medium.com/swlh/a-contemporary-introduction-to-natural-language-processing-nlp-in-2020-d93202502048. Published online May 10, 2020. Most recent access October 24, 2020.

13. Maini V. Machine learning for humans. https://medium.com/machine-learning-for-humans/why-machine-learning-matters-6164faf1df12. Published August 19, 2017. Parts 1 and 4 recommended. Accessed various times. Most recent access March 27, 2021.

14. The Cancer Genome Atlas. National Cancer Institute. https://www.cancer.gov/about-nci/organization/ccg/research/structural-genomics/tcga. Published 2008 with updates through 2018. Accessed various times. Most recent access October 26, 2020.

15. U.C. Berkeley School of Information. Story of big data, cancer and collaboration. https://dataedge.ischool.berkeley.edu/2017/schedule/story-big-data-cancer-and-collaboration. Published May 9, 2017. Last modified May 25, 2017. Accessed various times. Most recent access October 28, 2020.

16. Interpreting cancer genetic testing results. https://www.jax.org/education-and-learning/clinical-and-continuing-education/cancer-risk-assessment-testing-and-management/igtr. Accessed various times. Most recent access March 26, 2021.

17. Precision medicine for your practice: Interpreting results from somatic cancer panels. https://edhub.ama-assn.org/jackson-labs-learning/interactive/18113653. Accessed various times. Most recent access March 26, 2021.

18. Digital Health Becomes a Pillar: Tools, Payment and Data. Partners Healthcare World Innovation Summit. https://worldmedicalinnovation.org/past-forum-2020. Last Accessed March 27, 2021.

19. Lincoln SE, Nussbaum RI, Kurian AW, et al: Yield and utility of germline testing following tumor sequencing in patients with cancer. JAMA Network Open. 2020; 3 (10): e2019452. doi.10.1001/jamanetworkopen.2020.19452.

20. Wartman SA, Combs DC: Medical education must move from the information age to the age of artificial intelligence. Acad Med 2018 (8); 93: 1107–1109.

21. Densen P: Challenges and opportunities facing medical education. Trans Am Clin Climat Assoc 2011; 122: 48–58.

22. Shortliffe EH: Biomedical informatics in the education of physicians. JAMA 2010; 304 (11): 1227–1228.

23. Park SH, Do KH, Kim S, Park JH, Lim YS: What should medical students know about artificial intelligence in medicine? J Educ Eval for Health Prof 2019; 16: 18.

## Literature Citations / Commentary – MEDC-109 Spring Term 2021

1. Green ED, Guyer MS: Charting a course for genomic medicine from base pairs to bedside. Nature 2011; 470: 204–213. This 2011 paper provides an excellent summary. It may be supplemented by Green, Eric: The genomics landscape 2016. Current topics in genome analysis. https://www.genome.gov/event-calendar/Current-Topics-in-Genome-Analysis. The July 2018 video in this Eric Green Genomics Landscape group also is excellent.

2. Green ED, Rubin EM, Olson MV: The future of DNA sequencing. Nature 2017; 550: 179–181.

3. Wible B: Complicated legacies: The human genome at 20. Science 2021; 371: 564–569. This is a compendium of nine short essays by a variety of authors on various current topics in genomics. Of particular relevance to MEDC-109 are the essay on, “Algorithmic biology unleashed (H. Stevens, p. 565), “Value and affordability in precision medicine” (K. Phillips, J. Jansen, C. Weyant, p. 566) and “Genetic privacy in a post-COVID world” (D. Zielinski, Y. Erlich, pp. 566-567).

4. Hutson M: AI glossary; Artificial intelligence in so many words. Science 2017; 357: 19. (doi10.1126/science-357.6346.19) We will introduce this glossary in the opening session.

5. Miotto R, Wang F, Wang S, Jiang X, Dudley JT: Deep learning for healthcare: review, opportunities, challenges. Brief Bioinform 2018; 19(6): 1236–1246. (https://pubmed.ncbi.nlm.nih.gov/28481991/)

6. Esteva A, Robicquet A, Ramsundar B, et al: A guide to deep learning in healthcare. Nature Medicine 2019; 25: 24–29. (https://doi.org/10.108/s41591-018-0316-Z)

7. Hripcsak G, Freidman C, Alderson PO, et al: Unlocking clinical data from narrative reports: A study of natural language processing. Ann Int Med 1995; 122: 681–688.

8. Hirschberg J, Manning CD: Advances in natural language processing. Science 2015; 349: 261–266.

9. Hripcsak G, Albers DJ: Next generation phenotyping of electronic health records. JAMIA 2013; 20(1): 117–121. (NLP paper related to EHRs).

10. Roberts K, Boland MR, Pruinelli L … Brennan PF: Biomedical informatics advancing the national health agenda: the AMIA 2015 year-in-review in clinical and consumer informatics. JAMIA 2016; 24: e185–e190, https://doi.org/10.1093/jamia/ocw103. (the EHR section near the end provides good insights into how EHR’s can be improved).

11. Obermeyer Z, Emanuel EJ: Predicting the future - big data, machine learning and clinical medicine. N Engl J Med 2016; 375: 1216–1219.

12. Wartman SA, Combs DC: Medical education must move from the information age to the age of artificial intelligence. Acad Med 2018; 93(8): 1107–1109.

13. Harish V, Morgado F, Stern AD, Das S: Artificial intelligence and clinical decision making: The new nature of medical uncertainty. Acad Med 2021; 96(1) 31–36.

14. Hutson M: Bringing machine learning to the masses Science 2019; 365: 416-417. (introduces automatic machine learning – AutoML). (Supplement by reading about the Auto ML system being developed at MIT. http://news.mit.edu/2019/drag-drop-data-analytics-0627).

15. Rigby MJ: Ethical dimensions of using artificial intelligence in health care. AMA J Ethics 2019; 21: 121–124. doi 10.1001/amajethics.2019.121 (Try the several interesting hot links in the article to learn more. Also refer to website #11 in the listing that follows.)

## Websites / Commentary – MEDC-109 Spring Term 2021

1. OMIM (WWW.omim.org) This on-line compendium of human genes and genetic disorders was developed by and is curated daily at Johns Hopkins. It is based on the highly respected Johns Hopkins textbook “Mendelian Disorders in Man” published by Hopkins professor Victor McKusick in the 1960’s. Because this website was created from what was originally a book, the site contains excellent textual descriptions of thousands of disorders and links genotypes with phenotypes. It is an excellent site with which all first-year medical students should become familiar.

2. Gene Cards (www.genecards.org) The gene cards site was developed by and is curated at the Weizmann Institute of Science in Israel. It was started in 1997 and links to over 150 other websites. Disorders also can be searched and link to tabular presentations of genes associated with a disorder, function of genes, expression of the gene, etc. For the purposes of MEDC-109, the Gene Cards site is a detailed and worthy complement to OMIM. Genomic scientists might prefer Gene Cards.

3. Clinical Genomics Resource (www.clinicalgenome.org) The “ClinGen” resource was developed in 2013 by the National Human Genome Research Institute (NHGRI) at the NIH. It provides a central source where the clinical importance of numerous gene variants is defined to support precision medicine and research. The site addresses whether certain genes cause a clinical disorder and how the problems caused by variants might be treated.

4. “ClinVar” (www.ncbi.nlm.nih.gov/clinvar). This website is an NIH-curated source that was launched, like ClinGen, in 2013. It provides tabular listings of gene variants and disorders and can be searched using the disorders or gene symbols. We will learn to use ClinVar during this course.

5. Machine Learning for Humans – A beginners guide to AI/ML (www.medium.com/machine-learning-for-humans) This relatively brief clear English non-technical primer was written in 2017 by Vishal Maini, a Yale graduate who then was the communications manager for Deep Mind.com. The primer is divided into 8 parts, is well illustrated and contains many hot links for further exploration. At a minimum I suggest that you review Part 1 (“Why ML Matters”) and Part 4 (“Neural Networks and Deep Learning”).

6. Getting Started with NLP (Part 1) (www.medium.com/@gon.esboy/get-started-with-nlp) This is another introductory summary. It addresses natural language processing (NLP) in basic ways and provides links to more advanced study for those who are interested. Modern NLP makes great use of deep learning technologies, which is where this introduction is leading.

7. The Cancer Genome Atlas Program (tcga) (https://www.cancer.gov/about-nci/organization/ccg/research/structural-genomics/tcga) The data in this NIH/National Cancer Institute (NCI) program were compiled over 12 years. The site documents the genomic basis of 33 common cancers from over 11,00 patients. Twenty research sites collaborated with the NCI to create this rich database.

8. A Story of Big Data. Cancer and Collaboration. (https://dataedge.ischool.berkeley.edu/2017/schedule/story-big-data-cancer-and-collaboration) This is a 20-minute video about the collaboration of a patient – Shirley Pepke, a Ph.D. computational biologist, with Greg Ver Steeg, a Ph.D. physicist in computer science, who had developed a machine learning algorithm that allowed Pepke and Ver Steeg to explore the biology of Dr. Pepke’s own (previously) treatment resistant ovarian cancer. It is an excellent example of how the connecting genomics with machine learning can make a positive impact on patient care. It also is an excellent step-wise exploration of the issues encountered in analysis of complicated polygenic cancers. Dr Pepke’s cancer remains in remission.

9. Cancer variants database (www.civicdb.org) This database is curated at Washington University Medical School (WUMS) here in St. Louis. It is a good example of the importance of biobanking the myriad of gene variants that occur in cancer and how cancer-centric organizations share attributes of cancer variants. It contains a glossary, a clinical significance rating system and provides a portal to the Global Alliance for Genomics and Health, an international collaborative for sharing genomic information to benefit human health. Other organizations in this consortium include, for example, Memorial Sloan Kettering Cancer Center in NYC (www.oncokb.org).

10. Precision medicine for your practice: Interpreting results from somatic cancer panels. https://edhub.ama-assn.org/jackson-labs-learning/interactive/18113653 This is one of several excellent Jackson Lab educational sites that deal with how physicians might interact with patients who have questions raised by gene array testing.

11. How computerized facial recognition works (https://www.iflexion.com/blog/face-recognition-algorithms-work) This clearly written site shows the steps involved in computer vision analysis of facial features and, thus, personal identity. A site using a similar approach to render medical diagnoses can be found at www.face2gene.com.

12. “Ethics of artificial intelligence in healthcare and beyond”, Bander Center for Medical Business Ethics. P. Alderson and J. Kuene. (https://www.youtube.com/watch?v=bcUnfJ9_jTc) This session occurred in January of 2019 on SLU’s main campus. It is about 55 minutes long. The first four minutes are speaker introductions and Bander Center logistics, which can be skipped.

