## Supplementary material for "A Model to Introduce Medical Students to the Use of Artificial Intelligence and Genomics for Precision Medicine": Equator

### Equator – Criteria for Evaluating Training Interventions Healthcare

medRxiv

Set of reporting criteria for training interventions in health care. Van Hecke etal 2020 (#31 in list)

| N$^0$ | Criterion | Essential[a] (N) | Checklist |
|---|---|---|---|
| Development of the training | | | |
| 1 | Description of the aim or objectives of the training | 10 | ✓ |
| 2 | Description of the underlying theoretical framework | 3 | ✓ |
| 3 | Description of the developmental process | 8 | ✓ |
| 4 | Description of the target population and setting of the training | 9 | ✓ |
| 5 | Description of the educational resources | 9 | ✓ |
| Characteristics of the training | | | |
| 6 | Description of the content of the training | 9 | ✓ |
| 7 | Description of the format | 10 | ✓ |
| 8 | Description of the didactic methods of training | 8 | ✓ |
| 9 | Description of the tailoring of the training | 4 | ✓ |
| Characteristics of the providers/trainers | | | |
| 10 | Description of the provides of the training | 7 | ✓ |
| Assessment of the training outcomes | | | |
| 11 | Description of the measured outcomes | 9 | ✓ |
| 12 | Description of the applied assessment method, including validity and reliability | 9 | ✓ |

Authors: Alderson, Donlin, Morrison
